# The clinical impact of serious respiratory disease in children under the age of two during the 2021-2022 bronchiolitis season in the United Kingdom and Ireland

**DOI:** 10.1101/2023.02.22.23285616

**Authors:** Thomas C. Williams, Robin Marlow, Pia Hardelid, Mark D. Lyttle, Kate M. Lewis, Chengetai D. Mpamhanga, PERUKI, Steve Cunningham, Damian Roland

**Affiliations:** Department of Paediatric Respiratory and Sleep Medicine, Royal Hospital for Children and Young People, Edinburgh, UK; Child Life and Health, University of Edinburgh, UK; Emergency Department, Bristol Royal Hospital for Children, Bristol, UK; Great Ormond Street Institute of Child Health, University College London, UK; Research in Emergency Care Avon Collaborative Hub (REACH), University of the West of England, Bristol, UK; Centre for Inflammation Research, University of Edinburgh, UK; Paediatric Emergency Medicine Leicester Academic (PEMLA) Group, Leicester Royal Infirmary; Sapphire Group, Health Sciences, University of Leicester, UK

## Abstract

**Background:** Interventions introduced in 2020 to reduce the spread of SARS-CoV-2 led to a widespread reduction in childhood infections, including respiratory syncytial virus (RSV), in the subsequent year. However, from the spring of 2021 onwards the United Kingdom and Ireland began to experience an unusual out of season epidemic of childhood respiratory disease.

**Methods:** We conducted a prospective observational cohort study (BronchStart), enrolling children aged 0-23 months presenting with clinician-diagnosed bronchiolitis, lower respiratory tract infection or first episode of wheeze in 59 Emergency Departments (ED) across England, Scotland and Ireland from 1 May 2021 to 30 April 2022. We collected baseline data on patient demographics and clinical presentation, and follow-up data at 7 days. We used high-granularity BronchStart clinical data together with national English and Scottish admission datasets to infer the impact of RSV disease in a typical year before the Covid-19 pandemic, and to provide an up-to-date estimate of the annual impact of disease to inform implementation of anti-RSV interventions.

**Findings:** The BronchStart study collected data on 17,899 ED presentations for 17,179 children. Of these, 6,825 (38.1%) were admitted to hospital for further observation or treatment, 458 (2.6%) required care in a high dependency unit (HDU), and 154 (0.9%) were admitted to a paediatric intensive care unit (PICU). Of the 5,788 children admitted and tested for RSV, 41.8% of the overall study cohort, and 48.7% of those 0-11 months of age, were positive. Risk factors for hospital admission included prematurity and congenital cardiac disease. Patients with these risk factors were also more likely to receive oxygen therapy, or be admitted to a HDU or PICU. However, 84.5% of those admitted to an observation unit, 78.1% of those admitted to a ward, 67.7% of those admitted to HDU and 50.0 % of those admitted to PICU had no identified comorbidity. Using admissions data for England and Scotland we estimate that every year 12,167 infants with RSV infection receive low flow oxygen, 4,998 high flow oxygen and 6,198 a course of antibiotic therapy in secondary care.

**Interpretation:** Although RSV was the major pathogen in this cohort, 51.3% of admissions for serious respiratory viral infections in those aged <1 year of age were not associated with the virus. Whilst prematurity and congenital cardiac disease were risk factors for admission to hospital, HDU and PICU, the majority of these admissions, for all levels of care except PICU, were in previously healthy term born infants.

## Introduction

Respiratory Syncytial Virus (RSV) is an RNA virus that prior to the Covid-19 pandemic caused annual epidemics of respiratory infections, most prominently bronchiolitis. The main burden of bronchiolitis disease is in those under 1 year of age^1^. In temperate climates RSV epidemics, similar to influenza, normally occur in the autumn and winter^2^. Although RSV is the main driver of winter seasonal bronchiolitis epidemics, other viruses such as rhinovirus and human metapneumovirus are also important contributors. Globally, viral bronchiolitis is one of the largest contributors to hospital admission for children under the age of 5 years ^3–5^.

Non-pharmaceutical interventions, introduced in early 2020 to limit the spread of SARS-CoV-2, led initially to a reduction in the burden of all childhood infectious disease^6,7^ and subsequently to a disruption in the timing of annual RSV epidemics. Autumn and winter of 2021/2 saw very few RSV infections in children in Europe, the United States and China^8^. There was then an out of season increase in RSV in most of these countries the following spring or summer^9^, reflecting what had already been observed in the Southern Hemisphere^10^.

Despite significant investment, attempts to develop an effective RSV vaccine have proven unsuccessful until recentlyl^11,12^. Nirsevimab, a long-acting anti-RSV antibody targeting site 0 of the RSV fusion (F) protein, given as a single injection to infants at the start of their first RSV season, has recently shown significant benefit in a multinational randomised controlled trial^13^, with recent EMA and FDA licensing approval. There are similar promising results from a Phase III maternal immunisation study for RSV^14^. It is therefore likely that monoclonal antibodies or maternal immunisation will be introduced to routine clinical care soon, either for high-risk, or all, infants. Up-to-date information on the burden of disease is needed to inform the introduction of such immunisations.

The BronchStart study was initiated in the spring of 2021 to prospectively examine features of an anticipated unusual RSV summer season. At this point there were concerns that lack of exposure to viral infection in mothers and in the 0-23 month age group might lead to more severe disease and a shift in age distribution of disease, and we therefore focused the study on this population. We have previously reported our interim results, showing transposed seasonality but no evidence for increased severity of disease in this cohort^15^. Here, we present full data from the study, including hospital treatment burden in at-risk groups. Using the BronchStart cohort, we utilised national admissions data from England and Scotland from years before the Covid-19 pandemic to infer the total RSV-specific annual admissions and treatment burden for an average non-pandemic year, with the aim of informing decisions regarding the roll-out of any RSV preventative measure.

## Methods

### BronchStart data collection

BronchStart was a multi-national, multi-centre prospective observational study conducted at Paediatric Emergency Research in the UK and Ireland (PERUKI) Network sites^16^ supported by the RESCEU Consortium (www.resc-eu.org). The BronchStart protocol was structured in keeping with the principles of the STROBE statement^17^ and has been previously peer reviewed and published^18^.

Children aged 0-23 months presenting to participating Emergency Departments (EDs) with clinical features of bronchiolitis (cough, tachypnoea or chest recession, and wheeze or crackles on chest auscultation)^19^, lower respiratory tract infection or a first episode of acute viral wheeze were included in the study. Children with previous episodes of wheeze responsive to bronchodilator, suggesting an underlying diagnosis of recurrent wheeze of early childhood or early asthma, were excluded.

Data was collected on the date of attendance to a participating ED and seven days later. Anonymised data was entered onto a secure online database (REDCap data capture tool)^20,21^ hosted on a University Hospitals Bristol and Weston NHS Foundation Trust secure server. All data was collected by clinicians caring for children seen in the ED, or by clinical research teams based at the Trust where patients were seen. Data collected at baseline included patient demographics, comorbidities, presenting characteristics and disease acuity. The following comorbidities were collected in a structured format: preterm birth (gestational age <37 weeks), chronic lung disease of prematurity, congenital cardiac disease, and neuromuscular disorders; a free text box was also made available to enter additional comorbidities. An external link enabled clinicians to enter a full postcode derived index of multiple deprivation (IMD) score, a geographically based indicator of the relative deprivation of different areas. Data collected at 7 days included the child’s ultimate outcome (discharged or admitted), highest acuity dependency (the ward they were placed on if admitted: normal, high dependency or intensive care), respiratory support and pharmacological therapies administered in either the ED or whilst an inpatient, whether the patient was discharged or died and (if obtained) what viruses were identified by polymerase chain reaction (PCR) or other testing. RSV status was identified by nasopharyngeal aspirate (NPA) and (a) point of care testing (rapid viral testing where available) at baseline presentation to ED, and/or (b) by laboratory PCR testing, if either was performed as part of standard care. If a child tested positive using either of these, they were assumed to be RSV positive. The baseline and follow up questionnaires were externally peer reviewed and are publicly available^22^. Data was made available on a dashboard created by Microreact, in real-time^23^.

### National hospital admissions data

We used national datasets of admissions in England and Scotland for bronchiolitis, LRTI and wheeze for the years preceding the pandemic to estimate an annual national inpatient treatment burden. Due to the interruption to the end of the 2019-20 winter season by lockdown restrictions from February/March 2020 onwards, we used calendar years (January-December) to calculate a mean number of admissions in children 0-11 months, by year, from 2016 to 2019.

For England, we used Hospital Episode Statistics Admitted Patient Care (HES APC) data to identify admissions, as described previously^24^. HES APC is a database of all hospital inpatient admissions funded by the English National Health Service (NHS). Whilst it contains primary and secondary admissions codes, it does not contain information on treatments administered. For Scotland we used General Acute Inpatient and Day Case – Scottish Morbidity record (SMR01) data to identify admissions. SMR01 collects episode level data on hospital inpatient and day case discharges from acute specialties from hospitals in Scotland.

We identified all hospital admissions among children 0-23 months old (inclusive) for the period 1 January 2016 to 30 September 2022 with the following diagnoses: International Classification of Diseases version 10 (ICD-10) code J21 for bronchiolitis (primary or secondary diagnosis); J22 for lower respiratory tract infection (primary or secondary diagnosis); and viral wheeze (ICD-10 code B349 as primary diagnosis and R062 as any secondary diagnosis). We included multiple admissions per child but if a child had two or more of the above diagnoses recorded in the same admission, this admission was only counted once. Child’s sex, SIMD quintile (using doctor’s postcode) and age were also retrieved from the first episode in the patients admission.

### Outcomes

We considered the following outcomes for children with documented comorbidities: respiratory support provided (Oxygen [LF], Oxygen [HF], CPAP [continuous positive airway pressure]/BIPAP [bilevel positive airway pressure], IMV [invasive mechanical ventilation]) and admission to an HDU, (High Dependency Unit) or PICU (Paediatric Intensive Care Unit). For inference of national burden of disease, the interventions included were nasogastric fluids, intravenous fluids, low flow oxygen therapy, high flow oxygen therapy, non-invasive ventilatory support (Continuous Positive Airway Pressure [CPAP] or Bilevel Positive Airway Pressure [BiPAP]), and antibiotic administration. For antibiotic administration, antibiotics administered at any point during a child’s admission, including in the ED, were included.

### Statistical analyses

Data extraction and analysis was performed using RStudio and R version 4.2.2^25^. Descriptive tables were generated using gtsummary^26^. Risk ratios and confidence intervals for risk of different outcomes for infants born preterm (at <37 weeks) and/or with congenital cardiac disease were calculated using log binomial regression models. Data was also collected on chronic lung disease and neuromuscular disease; due to low numbers, risk ratios were not calculated for this group.

BronchStart is unique in joining up data on ED attendances, admissions, HDU and PICU admissions and secondary care prescribing, with information on PCR results for RSV testing from multiple sites across the UK. We used these data and a basic framework to infer the RSV specific hospital treatment burden for years preceding the pandemic. We first calculated the percentage of children testing positive for RSV in all patients who were tested for the virus in the BronchStart dataset, and applied these to the annual mean admission rate for all admissions in those aged 0-11 months in England (from the HES dataset) and Scotland (from the SMR01 dataset). The confintr^27^ package was used to calculate a two-sided 95% confidence interval for RSV positivity using the Clopper-Pearson confidence method, which was then applied for the proportion of the BronchStart admissions who received an intervention. Note that these analyses assume that the percentage of children admitted and who tested positive for RSV with particular diagnoses in the BronchStart dataset are representative of all children admitted to hospital for the relevant diagnoses in England and Scotland, and that incidence of each outcome in BronchStart was similar to that of England and Scotland in a pre-pandemic period. Analysis scripts are available at GitLab (https://git.ecdf.ed.ac.uk/twillia2/bronchstart-impact-of-rsv-analysis).

### Ethics

The study was initially exempted from ethical review in keeping with the Control of Patient Information Notice (COPI) under a Covid-19 Notice (Regulation 3(4) of the Health Service Control of Patient Information Regulations 2002) with University Hospitals of Leicester NHS Trust as the Study Sponsor. Following a query raised about the study being adopted onto the English National Institute for Health Research (NIHR) Clinical Research Network (CRN) portfolio the study was formally reviewed by the London City and East Ethics Committee and granted permission to recruit patients without formal consent (Reference 21/REC/1844). The study is registered on ClinicalTrials.gov (NCT04959734)^28^.

The use of Hospital Episode Statistics data was approved by the Health and Social Care Information Centre/NHS Digital (DARS-NIC-393510-D6H1D-v1.11). SMR01 data was provided by Public Health Scotland. Statistical disclosure control was applied to ensure patient confidentiality in line with the PHS Statistical Disclosure Protocol^29^.

### Public and patient involvement

The RSV Patient Advisory Board from ReSViNET^30^ were asked to comment on the initial BronchStart study design. As data were being collected anonymously, they agreed that it was acceptable to proceed without individual consent.

## Results

### Case recruitment to the BronchStart study

In the period 1 May 2021 to 30 April 2022, 17,935 ED attendances were recorded in the BronchStart study, of which, 17,899 attendances with complete entry were submitted for 17,179 individual children from 59 sites. Of the children, 15,933 (89%) were recruited at sites in England, 1,490 (8.3%) in Scotland and 476 (2.7%) in the Republic of Ireland (Supplementary Figure 1). Males predominated (10,839/17,899; 60.4%), and of 16,443 children with socio-economic status available, 2,701 (16.4%) were from the least deprived quintile, and 4,575 (27.8%) from the most deprived quintile.

Viral testing was most commonly performed for those admitted to hospital with RSV the most common single pathogen identified (Figure 1, panel A).Other viral pathogens were also identified in children tested, most commonly rhinovirus (929/17,899; 5.2%; Figure 1, panel B), where peaks in infection were similar in timing to RSV; peaks in RSV infection were followed temporally by spikes in human metapneumovirus (HMPV, 257/17,899;1.4%) and then SARS-CoV-2 (287/17,899;1.6%) infection.

**Figure 1.**
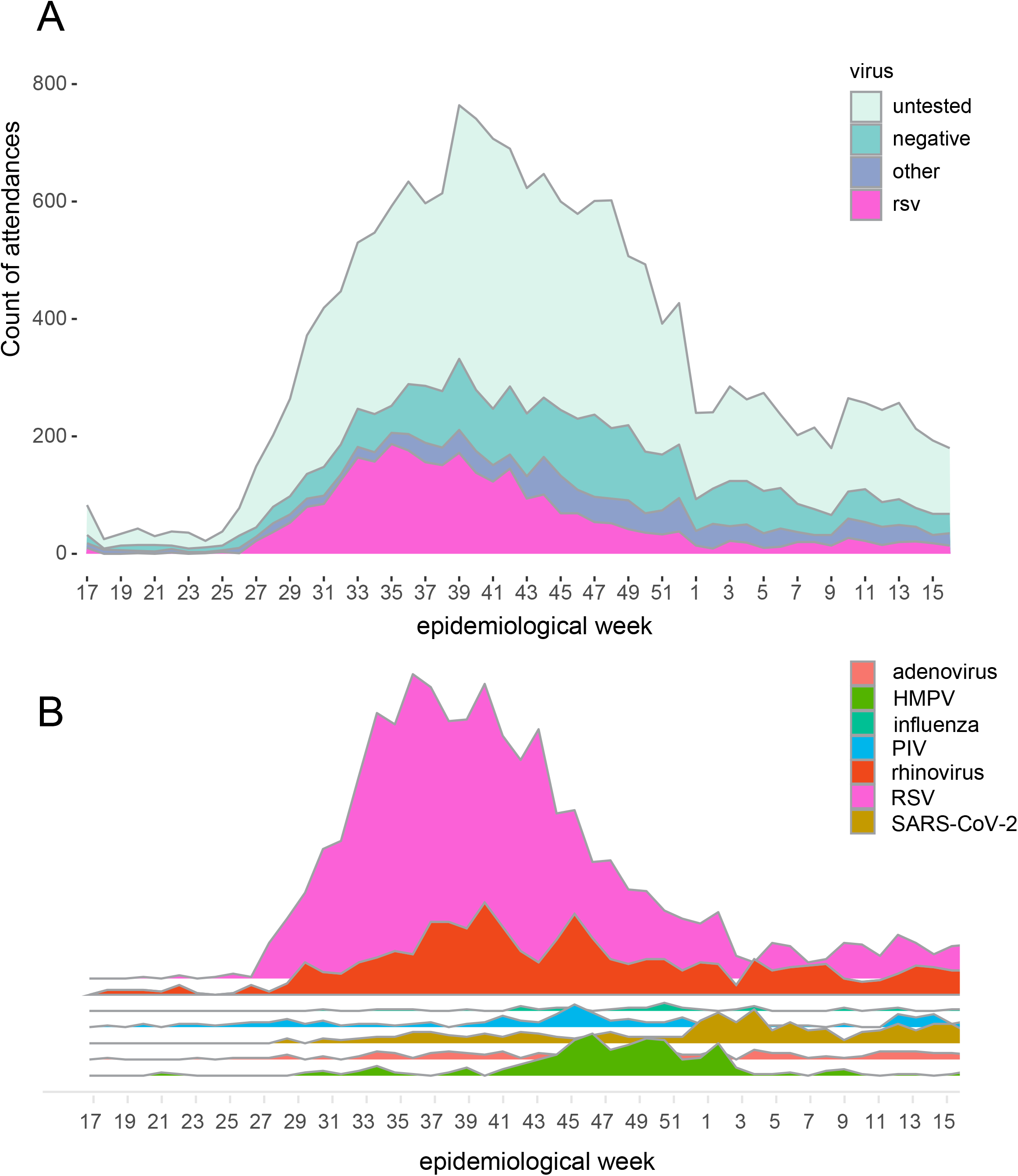
A) Virology testing showed RSV was the main pathogen during the study period. B) Graphical representation of other viruses identified in attendances in addition to RSV; area is proportional to number of positive tests. Abbreviations: HMPV (human metapneumovirus); PIV (Parainfluenza virus); SARS-CoV-2 (Severe acute respiratory syndrome coronavirus 2).

### Highest level of care

The most common diagnosis assigned to the BronchStart cohort was bronchiolitis (14,480/17,899; 80.9%), followed by first episode of viral induced wheeze (1,898/17,899; 10.6%) and lower respiratory tract infection (1,521/17,899;8.5%). The majority of children (11,074/17,899; 61.9%) were assessed in the ED (sometimes more than once) and discharged home (Table 1). A total of 6,825 (38.1%) children from the cohort were ultimately admitted to hospital; 2 children died in the ED. Again, the most common diagnosis assigned to those admitted was bronchiolitis (5,300/6,825;77.7%). The most common admission location was a hospital ward (5,027/6,825;73.7%) or an observation unit (1,186/6,825;17.4%). Of the entire BronchStart cohort, 458/17,899 (2.6%) required HDU level care, and a further 154 (0.9%) were admitted to a PICU (Table 1).

**Table 1.** Highest level of care provided to patients in the BronchStart cohort, by clinician diagnosis and diagnostic test and admission at day 7.

|  | ED,<br>N = 11,074 | Observation,<br>N = 1,186 | Ward,<br>N = 5,027 | HDU,<br>N = 458 | PICU,<br>N = 154 | Total,<br>N = 17,899 |
| --- | --- | --- | --- | --- | --- | --- |
| Diagnosis |  |  |  |  |  |  |
| Bronchiolitis | 9,180 (83%) | 847 (71%) | 3,918 (78%) | 396 (86%) | 139 (90%) | 14,480 |
| First Wheeze | 985 (8.9%) | 219 (18%) | 661 (13%) | 31 (6.8%) | 2 (1.3%) | 1,898 |
| LRTI | 909 (8.2%) | 120 (10%) | 448 (8.9%) | 31 (6.8%) | 13 (8.4%) | 1,521 |
| Virus |  |  |  |  |  |  |
| RSV | 431 (3.9%) | 189 (16%) | 1,951 (39%) | 195 (43%) | 85 (55%) | 2,851 |
| Other | 284 (2.6%) | 135 (11%) | 728 (14%) | 90 (20%) | 41 (27%) | 1,278 |
| Negative | 768 (6.9%) | 310 (26%) | 1,889 (38%) | 153 (33%) | 22 (14%) | 3,142 |
| Untested | 9,591 (87%) | 552 (47%) | 459 (9.1%) | 20 (4.4%) | 6 (3.9%) | 10,628 |
| Admission at day 7 |  |  |  |  |  |  |
| Still admitted at day 7 | 36 (0.3%) | 5 (0.4%) | 328 (6.5%) | 79 (17%) | 91 (59%) | 539 |
| Unknown | 109 (1.0%) | 0 (0%) | 1 (0.0%) | 0 (0%) | 0 (0%) | 110 |

Abbreviations: LRTI (Lower Respiratory Tract Infection); ED (Emergency Department); HDU (High Dependency Unit); PICU (Paediatric Intensive Care Unit); RSV (Respiratory Syncytial Virus).

Of 11,575 who attended the ED but were not immediately admitted (Supplementary Table 1), 9,928 were discharged and did not re-attend at 7 days, 969 re-attended but were discharged home again, and 678 re-attended and were admitted, meaning that after initial discharge there was a 14.2% (1,647/11,575) chance of re-attendance after initial discharge from an ED, and a 5.9% (678/11,575) chance of admission following initial discharge.

### Respiratory support and pharmacological therapy in the admitted BronchStart cohort

For the admitted cohort, low flow oxygen therapy was frequently administered for those with a diagnosis of bronchiolitis (2,310/5,301;43.6%), first episode of wheeze (301/913;33%) or LRTI (273/612;44.6%) (Table 2). High flow oxygen was administered most commonly for those with a diagnosis of bronchiolitis (922/5,301;17.4%), followed by those with LRTI (73/612;11.9%), and less commonly for those with a diagnosis of first episode of wheeze (47/913;5.1%). Antibiotics were administered to over two thirds of those with a diagnosis of lower respiratory tract infection (434/612;70.9%) and to over a fifth of those with a diagnosis of bronchiolitis (1,201/5,301;22.7%).

**Table 2:** Respiratory support and pharmacological therapies administered to admitted BronchStart patients, by clinician diagnosis.

| Treatment | Bronchiolitis, N = 5,301 | First Wheeze, N = 913 | LRTI, N = 612 |
| --- | --- | --- | --- |
| NG fluids | 1,975 (37%) | 30 (3.3%) | 62 (10%) |
| IV fluids | 686 (13%) | 39 (4.3%) | 105 (17%) |
| Oxygen (LF) | 2,310 (44%) | 301 (33%) | 273 (45%) |
| Oxygen (HF) | 922 (17%) | 47 (5.1%) | 73 (12%) |
| CPAP/BiPAP | 219 (4.1%) | 1 (0.1%) | 12 (2.0%) |
| IMV | 92 (1.7%) | 1 (0.1%) | 6 (1.0%) |
| Antibiotics | 1,201 (22.7%) | 157 (17.2%) | 434 (70.9%) |
| MgSO<sub>4</sub> | 27 (0.5%) | 37 (4.1%) | 9 (1.5%) |
| Prednisolone | 77 (1.5%) | 155 (17%) | 34 (5.6%) |
| Salbutamol inhaler | 781 (15%) | 784 (86%) | 249 (41%) |
| Salbutamol IV | 14 (0.3%) | 8 (0.9%) | 1 (0.2%) |

Abbreviations: LRTI (Lower Respiratory Tract Infection); NG (Nasogastric); IV (Intravenous); HF (High Flow); LF (Low Flow); CPAP (Continuous Positive Airway Pressure); BiPAP (Bilevel Positive Airway Pressure); IMV (Invasive mechanical ventilation); MgSO<sub>4</sub> (Magnesium Sulfate).

### Comorbidities in the BronchStart cohort

Of the BronchStart cohort, 3,011 (16.8%) were classified as having one or more comorbidities, including preterm birth. Of the cohort, 1,970 (11%) were born preterm, 183 (1%) had chronic lung disease of prematurity, 264 (1.5%) had congenital cardiac disease, and 22 (0.1%) had a neuromuscular disorder. The proportion of comorbidities increased with level of care received. Of those seen and discharged from an ED, 86.4% had no documented comorbidity; this fell to 50% of patients admitted to a PICU (Figure 2;Table 3). Prematurity was the most common comorbidity at all levels of care, including those admitted to HDU (89/458, 19.4%) and PICU (55/154, 35.7%). The most common comorbidity documented apart from prematurity or congenital heart disease in the HDU/PICU cohort was Down Syndrome (11/612 patients, 1.8%).

**Table 3.** Comorbidities and highest level of care received in the BronchStart cohort. Patients were ranked hierarchically so that each patient could only be assigned one category of comorbidity; congenital heart disease subsumed preterm birth or other comorbidities, and preterm birth other comorbidities.

| Highest level of care | CHD (%) | Preterm birth (%) | Other (%) | None (%) | Total |
| --- | --- | --- | --- | --- | --- |
| ED | 104 (0.9%) | 933 (8.4%) | 464 (4.2%) | 9,573 (86.4%) | 11,074 |
| Observation Unit | 19 (1.6%) | 113 (9.5%) | 52 (4.4%) | 1,002 (84.5%) | 1,186 |
| Ward | 109 (2.2%) | 699 (13.9%) | 293 (5.8%) | 3,926 (78.1%) | 5,027 |
| HDU | 23 (5.0%) | 89 (19.4%) | 36 (7.9%) | 310 (67.7%) | 458 |
| PICU | 9 (5.8%) | 55 (35.7%) | 13 (8.4%) | 77 (50.0%) | 154 |

Abbreviations: ED (Emergency Department); HDU (High Dependency Unit); PICU (Paediatric Intensive Care Unit); CHD (Congenital Heart Disease).

**Figure 2.**
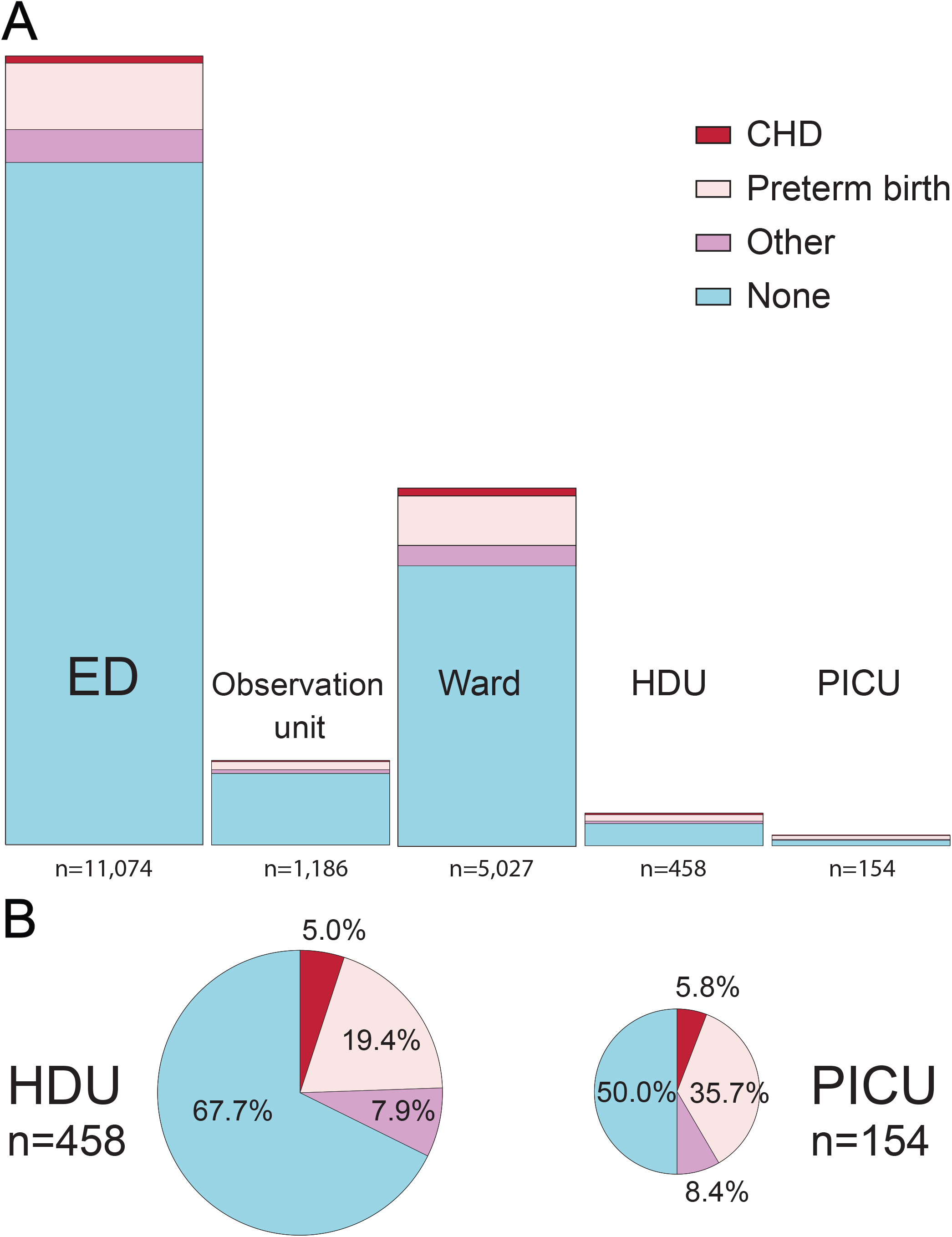
A) Highest level of care for BronchStart attendances including proportion of children with comorbidities. B) Children with comorbidities in HDU and PICU admissions. Data from Table 2, area of plots scaled to number of cases. Patients were ranked hierarchically so that each patient could only be assigned one category of comorbidity; congenital heart disease subsumed preterm birth or other comorbidities, and preterm birth other comorbidities. Abbreviations: CHD (Congenital Heart Disease); ED (Emergency Department); HDU (High Dependency Unit); PICU (Paediatric Intensive Care Unit).

### Burden of respiratory support for children with co-morbidities

Those born preterm had a 5.12-fold higher risk of invasive mechanical ventilation than those without comorbidities, and were more likely to be admitted, receive oxygen, receive high-flow oxygen therapy, be admitted to HDU/PICU, or receive invasive mechanical ventilation (Table 4). This higher risk persisted in participants aged 12-23 months (Supplementary Table 2).

**Table 4:** Treatments administered and highest level of care for those born preterm compared to those with no comorbidities in the BronchStart cohort.

| Outcome | No comorbidities (%)<br>N = 14,888 | Preterm (%)<br>N = 1,970 | RR | 95% CI |
| --- | --- | --- | --- | --- |
| Admitted | 5,316 (36%) | 1,007 (51%) | 1.43 | (1.36 – 1.5) |
| Oxygen (LF) | 2,262 (15%) | 528 (27%) | 1.76 | (1.62 – 1.91) |
| Oxygen (HF) | 754 (5.1%) | 221 (11%) | 2.22 | (1.92 – 2.55) |
| CPAP/BiPAP | 151 (1.0%) | 75 (3.8%) | 3.75 | (2.84 – 4.91) |
| IMV | 59 (0.4%) | 40 (2.0%) | 5.12 | (3.42 – 7.6) |
| Admit HDU | 327 (2.2%) | 117 (5.9%) | 2.70 | (2.19 – 3.31) |
| Admit PICU | 77 (0.5%) | 58 (2.9%) | 5.69 | (4.05 – 7.96) |

<sup>1</sup> Preterm group includes those who had associated comorbidities, aged 0-23 months.
<sup>2</sup> Relative risk and confidence intervals calculated using log binomial regression models.
Abbreviations: RR (Relative Risk); CI (Confidence Interval); HF (High Flow); LF (Low Flow); CPAP (Continuous Positive Airway Pressure); BiPAP (Bilevel Positive Airway Pressure); IMV (Invasive mechanical ventilation); HDU (High Dependency Unit); PICU (Paediatric Intensive Care Unit).

Participants with congenital cardiac disease were more likely to receive higher levels of care than those with no comorbidities (Supplementary Table 3), except for invasive mechanical ventilation, where although there was a high relative risk this did not reach statistical significance.

### Inferred national treatment burden

On average, there were 58,590 annual admissions for children in England and Scotland aged 0-11 months with a diagnosis of bronchiolitis, lower respiratory tract infection or wheeze in the period 2016-2019 (Supplementary Table 4). Comparing demographic variables for the BronchStart cohort to those for the HES and SMR01 admissions, age and socioeconomic were similar (Supplementary Table 5), but the BronchStart cohort had a higher proportion of infants in the 0-3 month age group (41.3% vs 32.3% in HES and 33.4% in SMR01). Of the admitted BronchStart cohort tested for RSV, 2,423/5,792 (41.8%) were positive RSV (1,907/3,912; 48.7% in those aged 0-11 months, 516/1880; 27.4% of those aged 12-23 months). Assuming an RSV positivity of 48.7% for the 0-11 month cohort gives an estimate of RSV positive admissions with these diagnoses per year for this age group. Using these estimates, we inferred that in an average year, RSV infections in England and Scotland were associated with 12,167 infants receiving low flow oxygen, 4,998 high flow oxygen, and 6,198 a course of antibiotics during their admission (Table 5).

**Table 5:** Inferred treatment burden for England and Scotland for bronchiolitis, lower respiratory tract infection and first episode of wheeze in infants 0-11 months. An annual admissions total was calculated for 2016-2019 using the HES (England) and SMR01 (Scotland) datasets.

| Treatment | BronchStart treatments | % treated in BronchStart cohort | % attributable to RSV | RSV specific treatments for annual admissions (95% CI) |
| --- | --- | --- | --- | --- |
| NG fluids | 1,884 | 41.6 | 20.3 | 11,881 (11,497-12,266) |
| IV fluids | 606 | 13.4 | 6.5 | 3,827 (3,703-3,951) |
| Oxygen (LF) | 1,925 | 42.6 | 20.8 | 12,167 (11,773-12,561) |
| Oxygen (HF) | 791 | 17.5 | 8.5 | 4,998 (4,836-5,160) |
| CPAP/BiPAP | 207 | 4.6 | 2.2 | 1,314 (1,271-1,356) |
| Antibiotics | 983 | 21.7 | 10.6 | 6,198 (5,997-6,399) |
| Total | 4,524 | 100 | 48.7 | 28,561 (27,637-29,486) |

Abbreviations: NG (Nasogastric); IV (Intravenous); LF (Low Flow); HF (High Flow); CPAP (Continuous Positive Airway Pressure); BiPAP (Bilevel Positive Airway Pressure); CI (confidence interval)

## Discussion

### Key results

The BronchStart study recruited 17,899 attendees with respiratory disease in under 2s, capturing information on comorbidities, respiratory support and pharmacological therapies administered. The most common diagnosis assigned was bronchiolitis, and RSV was the most common pathogen identified; however, only 48.7% of those admitted aged 0-11 months who were tested were positive for RSV. Risk factors for admission and escalation of therapy in the BronchStart cohort included prematurity and congenital heart disease. However, in our cohort for all levels of care, excluding PICU, most patients did not have any documented comorbidities. For PICU admissions 50% of children were documented as having comorbidities, of which preterm birth was the most common. Using the BronchStart dataset to infer the impact of disease, we confirmed that bronchiolitis and other respiratory diagnosis in infants under 1 year of age constitute a significant burden on secondary healthcare services.

### Strengths of the study

With BronchStart we have demonstrated the utility of rapidly rolled out research study to understand a novel presentation in children. The same research model is applicable to other conditions and other locations and could be rapidly implemented to understand changes in RSV and other respiratory disease following the introduction of any anti-RSV intervention.

Another strength of our study is the large number of cases recruited prospectively-we believe this is that largest prospective study of serious respiratory infections in children in the 0-23 month age group. We collected high granularity on viral testing, treatments administered and outcomes across EDs and hospital wards, at a point just before the likely introduction of RSV immunisation, and following the disruption caused by the COVID-19 pandemic. In addition, we estimate for the first time at a national level in England and Scotland the treatment, in addition to admission, burden of RSV specific respiratory disease in the 0-11 month age group.

### Limitations of the study

Recruitment to the study was limited to PERUKI study centres, and therefore we could not guarantee that the cases recruited were representative of the United Kingdom and Ireland as a whole. Comparison of BronchStart admissions to the HES and SMR-01 datasets for admissions with the same diagnoses showed that BronchStart admitted cases were more likely to be younger than admissions in England and Scotland in a pre-pandemic period.

Heterogeneity in testing approaches limits estimates of respiratory disease caused by other pathogens in this cohort. Our study does not include primary care prescription data, so our estimate relates solely to the antibiotic burden for RSV infection in admitted children. Another limitation is that we did not recruit children with suspected bacterial infection (sepsis, empyema) that may have developed secondary to RSV infection^31,32^, and thus we may have underestimated the burden of infection in this cohort. As the BronchStart study recruited patients following the lifting of COVID-19 restrictions, the percentage of cases positive for RSV, and the treatment burden, may not be representative of a typical year once RSV seasonality starts to resume normal seasonal patterns.

### Interpretation and comparison with other literature

We found that a lower proportion of admissions were positive for RSV than has been noted in previous studies^33,34^, which however focused on infants on infants and children with bronchiolitis rather than all serious respiratory viral infections. However, despite this lower proportion we estimate that in a typical year, a large number of infants with RSV infection in England and Scotland will receive low flow oxygen, high flow oxygen and a course of antibiotics. As well the direct costs of treatment, these have significant implications in terms of environmental impact^35^ and the costs of antimicrobial resistance^36^.

We found, as reported previously, that risk factors for admission and escalation of care in the BronchStart cohort included prematurity, congenital heart disease and other comorbidites^37,38^. Although part of this is likely to relate to national guidance^19^ to have lower admission thresholds for those with comorbidities, our dataset confirms the greater risk for those infants of requiring additional support, including critical care support.

Although we found a significant admission and treatment burden for RSV, our estimates vary considerably from recently published data. Using national surveillance data, Bardsley et al., ^39^ estimated a mean of 49,674 admissions for children under 1 year of age attributable to RSV disease in England in a typical winter in the period 2015-2019, which is almost double to what we estimate in our study for both England (population 56.5 million) and Scotland (population 5.4 million)^40^, and more than double than was estimated in a previous study^41^, albeit from a slightly earlier time period. Differences in our estimates may relate to the definition used in the study by Bardsley et al., where “RSV-associated disease presentations”^42^ were used rather than direct hospital based testing data, as used in this study, where results from both point of care testing and RT-PCR were used to infer an RSV-specific burden.

### Clinical and immunisation policy implications

We found that following discharge from an ED after presentation with symptoms of bronchiolitis, LRTI or wheeze, 14.2% of 0-23 month olds re-presented in the following 7 days, and 5.9% were admitted, which should help clinicians in counselling parents in this situation.

Our data suggest that even highly effective immunisation against RSV is likely to result in a reduction, rather than elimination, of seasonal admissions for respiratory viral infections in infants. This is because in our study only a proportion of serious respiratory viral presentations in infants were associated with RSV infection. Our study supports previous findings that comorbidities including those born preterm and congenital cardiac disease increase the probability of being admitted, of receiving treatment whilst an inpatient, and of escalation of care to HDU or PICU^43^. However, an immunisation approach that only targets high risk infants is unlikely to impact significantly on overall hospital burden, as we found that 77.9 % of admissions at all levels of care had no comorbidities.

### Conclusion

Our study provides an up-to-date estimate of the impact of acute respiratory disease due to viral infections in England and Scotland in those aged 0-23 months during a period of post-pandemic return to RSV circulation to guide immunisation decisions. Although RSV was the major pathogen in this cohort, over 50% of admissions for serious respiratory disease due to viral infection in those aged <1 year of age were not associated with the virus. Whilst prematurity and congenital cardiac disease were risk factors for admission to hospital, HDU and PICU, the majority of these admissions, for all levels of care, were in previously healthy term born infants.

## Data Availability

All data produced in the present study are available upon reasonable request to the authors

## Acknowledgments

We thank Khalil Abudahab, Anthony Underwood and David Aanenson at Microreact for support in creating the dashboard, and Linda Wijlaars for assistance in data extraction. We thank Mai Baquedano for technical support in the launch of the REDCap survey tool and ongoing data management, and Darren Goble for information management and technology support, including maintenance of the server and development of a data flow pipeline for the BronchStart outputs. We thank Elizabeth Whittaker for input at the project planning stage. We thank the RESCEU investigators for their support.

The use of Hospital Episode Statistics data was approved by the Health and Social Care Information Centre for the purpose of this study (DARS-NIC-393510-D6H1D-v1.11). Source data can be accessed by researchers applying to the Health and Social Care Information Centre for England. Copyright © 2018. Reused with the permission of the Health and Social Care Information Centre. All rights reserved. Research at UCL Great Ormond Street Institute of Child Health is supported by the NIHR Great Ormond Street Hospital Biomedical Research Centre This research benefits from and contributes to the NIHR Children and Families Policy Research Unit, but was not commissioned by the NIHR Policy Research Programme. This work uses data provided by patients and collected by the English NHS as part of their care and support.

## Supplementary Tables

**Supplementary Table 1:** Discharge pathways for patients in the BronchStart cohort.

| Attendance/Discharge pathway | Number | Percent of total cohort |
| --- | --- | --- |
| Dc | 9,928 | 55.5% |
| Dc/Dc | 969 | 5.4% |
| Dc/Ad | 678 | 3.8% |
| Ad | 333 | 1.9% |
| Ad/Dc | 5,371 | 30.0% |
| Ad/Dc/Dc | 198 | 1.1% |
| Ad/Dc/Ad | 246 | 1.4% |
| other | 77 | 0.4% |
| NA | 99 | 0.6% |

Abbreviations: Ad (admitted); Dc (discharged).

**Supplementary Table 2:** Outcomes for preterm patients aged 12-23 months in the BronchStart cohort, compared to those with no known comorbidities.

| Characteristic | No comorbidities,<br>N = 4,384 | Preterm,<br>N = 532 | RR | 95% CI |
| --- | --- | --- | --- | --- |
| Admitted | 1,810 (41%) | 291 (55%) | 1.32 | (1.21 - 1.44) |
| Oxygen (LF) | 761 (17%) | 167 (31%) | 1.81 | (1.56 - 2.08) |
| Oxygen (HF) | 166 (3.8%) | 60 (11%) | 2.98 | (2.23 - 3.92) |
| CPAP/BiPAP | 9 (0.2%) | 10 (1.9%) | 9.16 | (3.71 – 22.95) |
| IMV | 5 (0.1%) | 5 (0.9%) | 8.24 | (2.3 - 29.54) |
| Admit HDU | 73 (1.7%) | 24 (4.5%) | 2.71 | (1.69 - 4.19) |
| Admit PICU | 6 (0.1%) | 9 (1.7%) | 12.4 | (4.48 – 36.75) |

<sup>1</sup> Preterm group includes those who were born preterm and had associated comorbidities .
<sup>2</sup> Relative risk and confidence intervals calculated using log binomial regression models.
Abbreviations: RR (Relative Risk); CI (Confidence Interval); HF (High Flow); LF (Low Flow); CPAP (Continuous Positive Airway Pressure); BiPAP (Bilevel Positive Airway Pressure); IMV (Invasive mechanical ventilation); HDU (High Dependency Unit); PICU (Paediatric Intensive Care Unit).

**Supplementary Table 3:** Treatments administered and highest level of care for those with congenital heart disease compared to those with no comorbidities in the BronchStart cohort.

| **Outcome** | **No comorbidities (%)**<br>N = 14,888 | **CHD (%)**<br>N = 264 | **RR** | **95% CI** |
| --- | --- | --- | --- | --- |
| Admitted | 5,316 (36%) | 160 (61%) | 1.70 | (1.53 - 1.86) |
| Oxygen (LF) | 2,262 (15%) | 90 (34%) | 2.24 | (1.87 - 2.64) |
| Oxygen (HF) | 754 (5.1%) | 36 (14%) | 2.69 | (1.93 - 3.61) |
| CPAP/BiPAP | 151 (1.0%) | 15 (5.7%) | 5.60 | (3.2 – 9.05) |
| IMV | 59 (0.4%) | 3 (1.1%) | 2.87 | (0.7 - 7.67) |
| Admit HDU | 327 (2.2%) | 25 (9.5%) | 4.31 | (2.85 - 6.2) |
| Admit PICU | 77 (0.5%) | 9 (3.4%) | 6.59 | (3.1 - 12.28) |

<sup>1</sup> CHD group includes those who had associated comorbidities, aged 0-23 months.
<sup>2</sup> Relative risk and confidence intervals using log binomial regression models.
Abbreviations: RR (Relative Risk); CI (Confidence Interval); HF (High Flow) ; LF (Low Flow); CPAP (Continuous Positive Airway Pressure); BiPAP (Bilevel Positive Airway Pressure); IMV (Invasive mechanical ventilation); HDU (High Dependency Unit); PICU (Paediatric Intensive Care Unit).

**Supplementary Table 4.** Mean admissions for those aged 0-11 and 12-23 months in the HES (England) and SMR-01 (Scotland) datasets by age band and diagnosis for years 2016-2019, for the diagnoses of bronchiolitis, LRTI and viral wheeze. Each admission was counted for a patient only once.

|  | 2016 | 2017 | 2018 | 2019 | Mean |
| --- | --- | --- | --- | --- | --- |
| **England** |  |  |  |  |  |
| 0-11 months | 54,396 | 50,583 | 55,914 | 55,162 | 54,014 |
| 12-23 months | 33,733 | 32,986 | 36,072 | 36,279 | 34,768 |
| **Scotland** |  |  |  |  |  |
| 0-11 months | 4,739 | 4,256 | 4,359 | 4,949 | 4,575 |
| 12-23 months | 2,420 | 2,250 | 2,371 | 2,779 | 2,455 |
| **Total** |  |  |  |  |  |
| 0-11 months | 59,135 | 54,839 | 60,273 | 60,111 | 58,590 |
| 12-23 months | 36,153 | 35,236 | 38,443 | 39,058 | 37,223 |

Abbreviations: LRTI (lower respiratory tract infection).

**Supplementary Table 5:** Comparison of demographic variables for BronchStart and HES datasets in the 0-11 month age group. Q1 is the least deprived, and Q5 the most deprived.

| Cohort | Sex |  | IMD quintile |  |  |  |  | Age band |  |  |  |
| --- | --- | --- | --- | --- | --- | --- | --- | --- | --- | --- | --- |
|  | Female | Male | % Q1 | % Q2 | % Q3 | % Q4 | % Q5 | 0-<3m | 3-<6m | 6-<9m | 9-<12m |
| HES (England) | 38.0% | 62.0% | 14.7% | 15.2% | 17.6% | 21.6% | 30.9% | 32.3% | 26.1% | 21.9% | 19.8% |
| SMR01 (Scotland) | 39.2% | 60.8% | 13.2% | 17.2% | 16.7% | 22.5% | 30.4% | 33.4% | 26.8% | 21.8% | 18.0% |
| BronchStart | 40.2% | 59.8% | 17.0% | 16.7% | 17.3% | 20.9% | 28.1% | 41.3% | 24.4% | 18.1% | 16.2% |

Abbreviations: HES (Hospital Episode Statistics); IMD (index of multiple deprivation); Q (Quintile).

