## Supplementary Figure 1 for "The clinical impact of serious respiratory disease in children under the age of two during the 2021-2022 bronchiolitis season in the United Kingdom and Ireland"

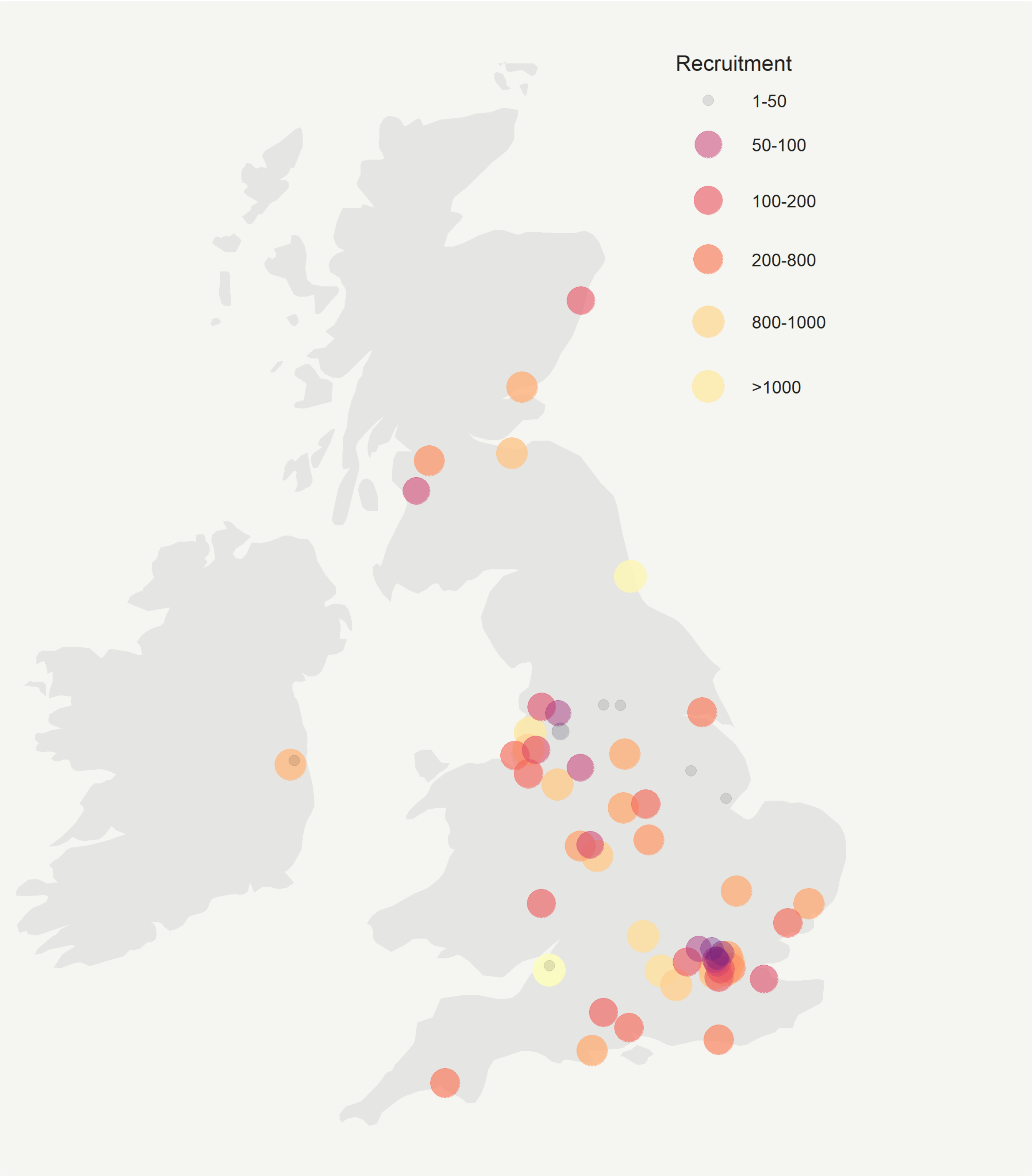


**Supplementary Figure 1   Recruitment to the BronchStart study from 1 May 2021 to 30 April 2022 by site location and number of attendance per site.**
